# Global co-hotspots of dengue, malaria, and yellow fever: a comparative spatiotemporal analysis across 142 countries

**DOI:** 10.64898/2026.07.11.26357638

**Authors:** Qiancheng Ma, Tianzhen Zhang

## Abstract

Dengue, malaria, and yellow fever remain major mosquito-borne public health threats in tropical and subtropical regions. Yet most global spatial studies have focused on single diseases or on potential transmission suitability rather than empirically observed shared burden. We therefore sought to identify robust global co-hotspots of these three diseases across 142 countries from 1990 to 2023, treating mosquito-borne diseases as an integrated public health problem. Using annual country-level incidence data from the Global Burden of Disease study and demographic, health-system, and climate covariates, we compared a fully Bayesian shared-component model, a neural network model with spatial-lag features, and a two-stage hybrid model. Shared scores were standardised for cross-model comparison and externally validated against an independent mortality anchor.

Overall, 29 countries were classified as consensus hotspots, all in sub-Saharan Africa, while seven countries formed a moderate-agreement watch list at the margins of the main hotspot belt, suggesting potential transition zones of shared burden.

By shifting the analytical focus from potential suitability to the empirical identification of realised burden and shared control vulnerability, this study provides actionable global evidence on where mosquito-borne disease prevention and control may be most structurally constrained. The findings may support more integrated prioritisation of vector control, surveillance, and health-system preparedness across countries.

## 1. Introduction

Dengue, malaria, and yellow fever continue to impose substantial public health burdens in many tropical and subtropical countries^1-3^. Although these diseases differ in aetiology and transmission ecology, they are shaped by overlapping ecological and climatic conditions, favourable mosquito habitats, and structural vulnerabilities in vector surveillance and control^4^.

Global spatial research on mosquito-borne diseases has developed along two main directions. One direction consists of numerous single-disease or region-specific spatial studies, together with systematic reviews that synthesise evidence from across the global literature^5^. Although this body of work has broadened our understanding of spatial approaches in mosquito-borne disease research, it is not designed to directly quantify the realised burden of individual countries or to identify where shared vulnerability in surveillance, vector control, and health-system preparedness is most concentrated across multiple diseases. A second direction is represented by global mapping studies that estimate environmental suitability, potential transmission conditions, or surveillance-adjusted risk. While such work is valuable for identifying where transmission may occur, it primarily addresses potential distribution rather than the sustained realised burden that multiple mosquito-borne diseases may jointly produce at the country level^6^. From a public health perspective, jointly examining dengue, malaria, and yellow fever may therefore provide a more actionable basis for identifying places where vector control, surveillance capacity, and health-system preparedness are simultaneously strained. In this context, our study asks a different question: which countries emerge as robust shared burden hotspots across dengue, malaria, and yellow fever, and do these priority zones remain stable across Bayesian, neural-network, and hybrid spatiotemporal frameworks?

Taken together, an important unmet need is to treat mosquito-borne diseases as a broader integrated public health problem and to empirically identify, on the basis of observed epidemiological burden, where shared control vulnerability is most concentrated at the global level. This study was designed to address that need.

## 2. Materials and methods

### 2.1. Statistical software

Spatial data processing and map production were conducted using ArcGIS Pro version 3.5.2 (Esri, Redlands, CA, USA). Additional data processing and modelling were conducted in Python version 3.13.13 (Python Software Foundation, Wilmington, DE, USA).

### 2.2. Data source and study scope

Data were obtained from the Global Burden of Disease (GBD) study. Countries with no reported cases of dengue, malaria, or yellow fever throughout the study period were excluded. The final dataset included 142 countries spanning 1990 to 2023.

Among them, dengue involved 126 countries with a total of 371 937 600 incident cases, malaria involved 104 countries with a total of 8 237 682 000 incident cases, and yellow fever involved 47 countries with a total of 6 232 417 incident cases. These country-level annual incidence and mortality data were used to construct the panel dataset for subsequent modelling. Temporal curves for the three diseases are shown in Figure 1.

**Figure 1.**
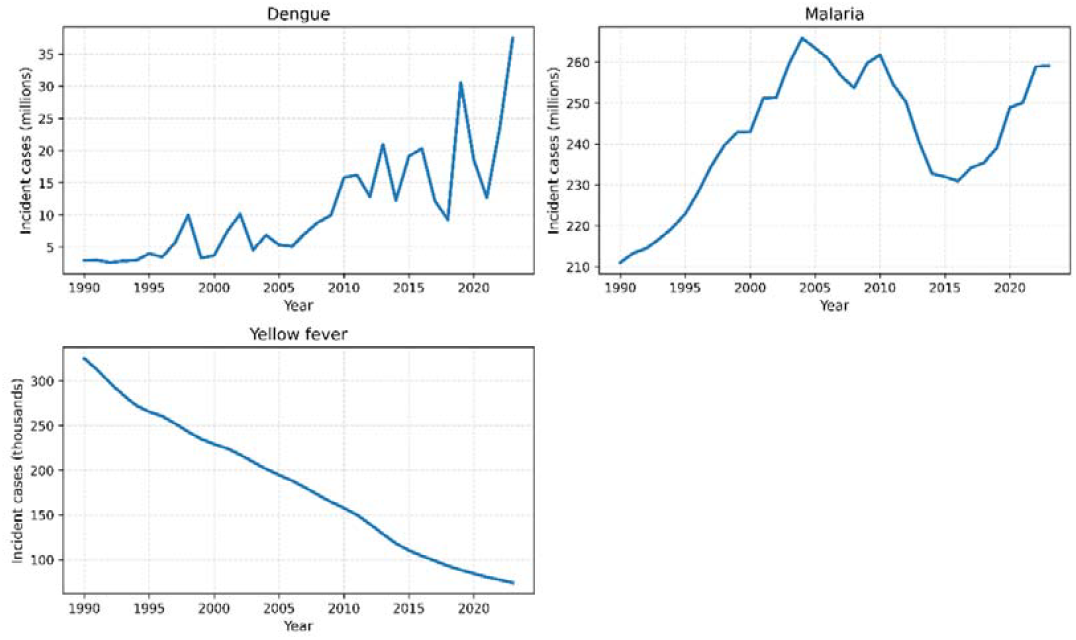
Temporal trends in incident cases of dengue, malaria, and yellow fever, 1990–2023 (Alt text: Line chart of annual incident cases of dengue, malaria, and yellow fever from 1990 to 2023.)

### 2.3. Statistical models

Detailed information can be found in Supplementary Material Text 1.

We compared three spatiotemporal modelling paradigms using the same country-year panel of 142 countries observed from 1990 to 2023 for three mosquito-borne diseases: dengue, malaria, and yellow fever. Let denote the observed case count for country, disease, and year, with population offset . The detailed data workflow of this study is shown in Figure 2.

**Figure 2.**
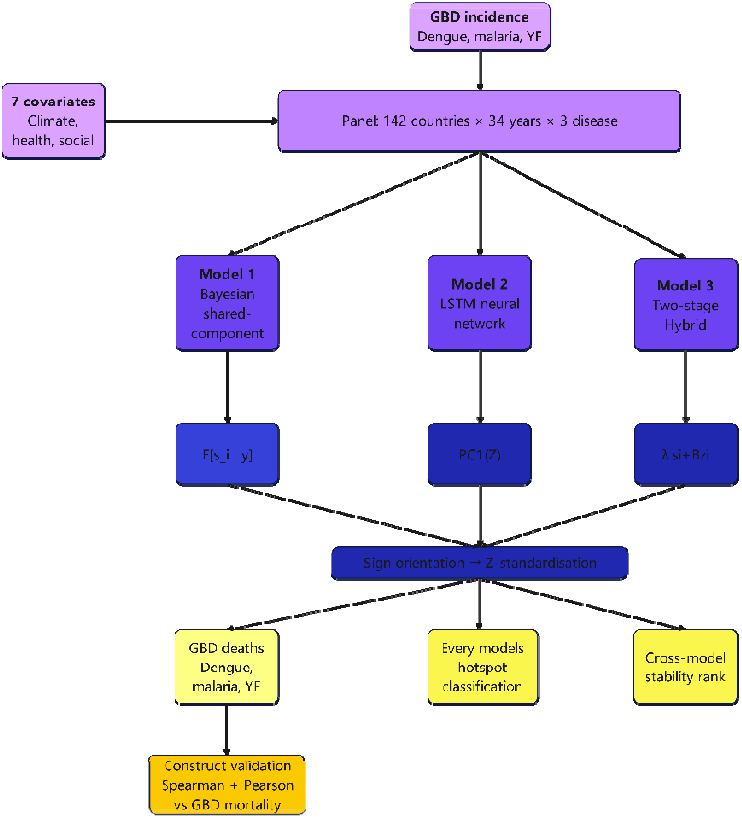
Study data and analytical workflow (Alt text: Flowchart of the study design, data sources, modelling steps, and validation process.)

#### 2.3.1. Model 1: Fully Bayesian shared-component model

We fitted a hierarchical Negative Binomial shared-component model jointly across the three diseases to capture their residual common geographic risk structure after accounting for covariates, disease-specific spatial variation, and temporal trends^7,8^.

The observation model was specified as:

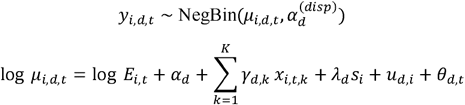

Where:

*μ*_*i,d,t*_ is the expected case count for country *i*, disease *d*, and year *t*;

*E*_*i,t*_ is the population offset; *α*_*d*_ is the disease-specific intercept;

*γ*_*d,k*_ is the disease-specific coefficient for covariate *k*;

*x*_*i,t,k*_ is the standardised value of covariate *k* for country *i* in year *t*;

*λ*_*d*_ is the disease-specific loading on the shared spatial component;

*s*_*i*_ is the latent shared spatial component;

*u*_*d,i*_ is the disease-specific residual spatial effect;

*θ*_*d,t*_ is the disease-specific first-order random-walk [RW(1)] temporal trend.

**The primary reported output** from Model 1 was the continuous shared spatial score, defined as the posterior mean of the latent shared component for each country, 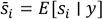 . This quantity was interpreted as each country’s posterior mean contribution to the geographic risk structure shared across the three diseases.

##### Diagnostics

Posterior convergence was assessed using the potential scale reduction factor 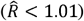 and effective sample size (ESS), supplemented by posterior predictive checks on the log-count scale^9,10^. Model fit was summarised using widely applicable information criterion (WAIC) and leave-one-out cross-validation (LOO), with comparisons restricted to within-family interpretation because the likelihood structures differed across the three modelling paradigms^11^.

All unknown parameters and latent effects were assigned prior distributions and estimated jointly under a Bayesian framework. In particular, the shared spatial component, disease-specific residual spatial effects, temporal random-walk terms, regression coefficients, and overdispersion parameters were inferred from their joint posterior distribution using Markov chain Monte Carlo sampling. Full prior specifications are provided in the Supplementary Text 1.

#### 2.3.2. Model 2: Neural network (NN)

A two-layer LSTM encoder with three disease-specific regression heads was trained on the same country × year × covariate panel, augmented with row-normalised spatial-lag features to ensure fair comparison with the Bayesian model^12^:

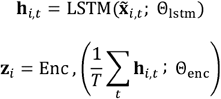

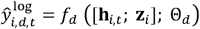

**where:**

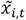: concatenation of standardised country-level covariates and their spatial lag *W*·*x*, with *W* the row-normalised adjacency matrix

*h*_*i,t*_ ∈ ℝ^*H*^: hidden state from a 2-layer LSTM (*H* = NN_HIDDEN)

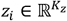: time-aggregated country-level latent embedding produced by a small MLP encoder (*K*_z_= NN_EMBED)

*f*_*d*_: disease-specific feed-forward head mapping [*h*_*i,t*_ ; *z*_*i*_]to the predicted log incidence rate *log(cases/pop × 10*^*5*^*+ 1)*

The network was trained to minimise mean squared error (MSE) on the log-rate outcome using the Adam optimizer, with weight decay, gradient clipping, learning-rate scheduling, and early stopping based on a held-out validation subset of the training years.

**The Neural network -derived shared score** is defined as the first principal component of the country-level embeddings *z*_*i*_, sign-oriented to align with observed log mortality.

The model was trained on data from 1990 to 2019, with the final 20% of training years used as a validation subset for early stopping and hyperparameter control. Predictive performance was then evaluated on held-out test years from 2020 to 2023.

##### Diagnostics

(i) training/validation MSE trajectories and early-stopping epoch; (ii) out-of-sample test MSE on held-out years (post-TRAIN_END); (iii) predicted-vs-observed scatter by disease on the log-rate scale.

#### 2.3.3. Model 3: Two-stage Hybrid (Neural network representation + Bayesian observation layer)

The Hybrid model uses the country-level latent embeddings from Model 2 as fixed covariates in a Bayesian observation layer. This second-stage model re-estimates the shared spatial structure, disease-specific temporal trends, and uncertainty while preserving the nonlinear country representation learned by the neural network.

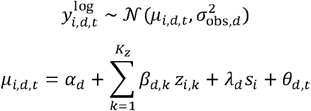

where

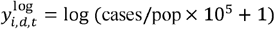 is the log-incidence-rate outcome for country *i*, disease *d*, and year *t*; *α*_*d*_ is the disease-specific intercept;

*β*_*d,k*_ is the disease-specific coefficient on the *k*-th NN latent feature;

*z*_*i,k*_ is the *k*-th component of the fixed NN country embedding imported from Model 2;

*λ*_*d*_ is the disease-specific loading on the shared spatial component;

*s*_*i*_ is the shared scaled-ICAR spatial field;

*θ*_*d,t*_ is the disease-specific first-order random walk (RW(1)) temporal trend. The observation-level residual variation was modelled through *σ*_obs,*d*_, with disease-specific standard deviations.

In the second stage, all unknown Bayesian-layer parameters and latent effects were assigned prior distributions and estimated jointly from their posterior distribution; detailed prior specifications and implementation are provided in the Supplementary Text 1.

##### Rationale

The two-stage design deliberately separates representation learning (where NN flexibility is useful) from structural inference (where Bayesian machinery is needed). This avoids the identifiability conflict we observed in earlier pilots when NN latent features and Bayesian spatial/temporal components were estimated jointly.

##### Diagnostics

(i) 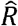 and ESS for *β*_*d,k*_, *λ*_*d*_, *σ*_*s*_, *σ*_*rw,d*_, and *σ*_*obs,d*_ ; (ii) divergent transitions; (iii) posterior predictive checks on the log-rate scale; and (iv) within-model WAIC and LOO.

#### 2.3.4. Shared score, hotspot classification, and cross-model stability

For each model *m* ∈ *Bayes, NN, Hybrid*, the country-level shared score 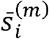 was sign-oriented against observed disease anchors to ensure comparability. Scores were standardised to Z-scores, and countries with 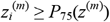 were labelled as hotspots under model *m*, where *P*_75_ denotes the 75th percentile of the standardised score distribution across 142 countries. The cross-model stability rank was defined 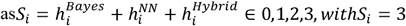 denoting consensus hotspots.

#### 2.3.5. External validation

All three shared scores were validated against the GBD cumulative log-mortality rate for the same three diseases (aggregated over the entire study period, 1990–2023) using Spearman and Pearson correlations across the 142 countries.

#### 2.3.6. Covariates and data sources

The covariates included population density, urbanisation rate, physician density, the universal health coverage (UHC) index, temperature, humidity, and rainfall; all were obtained from World Bank data sources, with the climate variables derived from the World Bank Climate Change Knowledge Portal.

## 3. Results

### 3.1. Model diagnostics and convergence

Detailed model diagnostic information and interpretation of the results are provided in Supplementary Text 2.

#### 3.1.1. Bayesian shared-component diagnostics

Model 1 showed satisfactory convergence and fit, with all 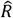 values below 1.01, minimum *ESS*_*bulk*_ and *ESS*_*tail*_ of 492.4 and 1050.2, no post-tuning divergences, acceptable posterior predictive fit, and broadly concordant WAIC and LOO values.

#### 3.1.2. Neural network (LSTM + spatial lag) diagnostics

Model 2 (NN) was successfully trained under an explicit early-stopping rule. Training and validation MSE both decreased substantially, although the later plateau in validation loss suggested mild overfitting. Observed-versus-predicted plots showed positive correspondence across all three diseases, but with compressed predictions and systematic underestimation at higher observed log-rates, indicating weaker fit in the upper tail.

#### 3.1.3. Two-stage Hybrid diagnostics

Model 3 (Hybrid) showed acceptable convergence and fit, with all 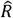 values below 1.01, minimum *ESS*_*bulk*_ and *ESS*_*tail*_ of 229.2 and 533.2, no post-tuning divergences, acceptable posterior predictive fit on the log-rate scale, and broadly concordant WAIC and LOO values within the Gaussian-on-log-rate family.

### 3.2. External construct validation

Three models showed strong and statistically significant positive associations with the GBD cumulative log-mortality anchor across 142 countries. The Bayesian model achieved the highest Spearman correlation (*ρ*_*S*_ = 0.837), indicating the best agreement with the external benchmark in terms of country ranking. In contrast, the Hybrid model achieved the highest Pearson correlation (*ρ*_*P*_ = 0.829), narrowly exceeding the NN model (*ρ*_*P*_ = 0.828), suggesting the strongest linear correspondence with the external mortality burden. These results indicate a complementary pattern across models: the Bayesian model was most effective for preserving relative country ordering, whereas the Hybrid model was most effective for capturing the overall linear gradient of external disease burden. All correlations were highly significant (*p* < 10^−30^).

### 3.3. Shared hotspot scores and geographic patterns

Detailed data are presented in Supplementary Table S1.

#### 3.3.1. Three-model shared-score maps

Using the Z-standardised shared scores, all three models identified sub-Saharan Africa (SSA) as the dominant high-risk region for mosquito-borne disease co-occurrence. In all three maps, the highest positive scores were concentrated primarily in equatorial and tropical parts of West, Central, and East Africa. Outside SSA, most countries showed low or negative standardised scores, including North Africa (e.g., Egypt), the Middle East (e.g., Kuwait, Lebanon, Jordan, Saudi Arabia), Central Asia (e.g., Georgia), East Asia (e.g., China and the Republic of Korea), and several Pacific Island settings such as the Cook Islands. Figure 3 presents the standardised shared-score maps for the Bayesian, neural network, and Hybrid models.

**Figure 3.**
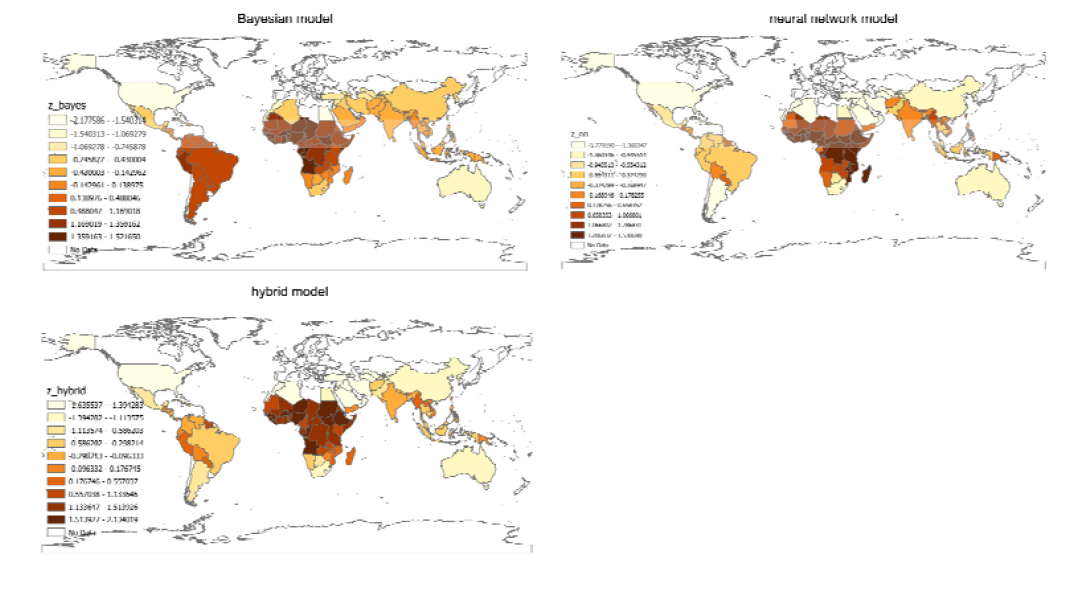
Shared hotspot score maps from the Bayesian, neural network, and Hybrid models (Alt text: Three world maps showing shared hotspot scores from the Bayesian, neural network, and hybrid models.)

#### 3.3.2. Bayesian shared-score map

High scores were concentrated in West and Central Africa, with visible within-SSA gradation, such that neighbouring countries often showed distinct score levels rather than forming a single homogeneous hotspot block. The top five countries were Liberia (+1.52), Nigeria (+1.50), Gabon (+1.48), Congo (+1.45), and Burkina Faso (+1.45). The lowest-scoring countries were the United States (−2.20), Egypt (−1.85), the Cook Islands (−1.68), Lebanon (−1.64), and Palestine (−1.53).

#### 3.3.3. NN shared-score map

SSA countries formed a more spatially coherent high-score block, with less internal gradation than in the Bayesian map. The top five countries were Guinea (+1.54), Sierra Leone (+1.52), Guinea-Bissau (+1.51), Togo (+1.51), and Uganda (+1.50). Several East and Southern African countries also received high NN scores, including Madagascar (+1.43), Mozambique (+1.43), Malawi (+1.40), and Comoros (+1.29). The lowest-scoring countries were Kuwait (−1.78), Lebanon (−1.77), Jordan (−1.74), Saudi Arabia (−1.69), and Georgia (−1.64).

#### 3.3.4. Hybrid shared-score map

The Hybrid model combined broad SSA spatial coherence with sharper internal differentiation. Its top five countries were Liberia (+2.13), Burundi (+1.92), Guinea (+1.91), Sierra Leone (+1.90), and South Sudan (+1.83). The lowest-scoring countries were Kuwait (−1.64), Lebanon (−1.63), Jordan (−1.62), Saudi Arabia (−1.60), and Georgia (−1.57). Among the three models, the Hybrid map showed the strongest positive separation at the upper end of the standardised score distribution.

### 3.4. Cross-model comparison of hotspot and coldspot patterns

#### 3.4.1. Hotspot agreement

All three models placed their top five countries exclusively in SSA, but the specific rankings differed. No country appeared in the top five of all three models simultaneously. Liberia appeared in the top five of the Bayesian and Hybrid models, but not the NN model. Guinea and Sierra Leone appeared in the top five of the NN and Hybrid models, but not the Bayesian model. The Bayesian and NN top-five sets had no overlap.

One of the clearest geographic disagreements concerned East and Southern Africa. Madagascar (+1.43), Mozambique (+1.43), Malawi (+1.40), and Comoros (+1.29) all received high NN scores, but their Bayesian scores were much lower (Madagascar −0.02, Mozambique +0.14, Malawi +0.19, and Comoros +0.25). Their Hybrid scores were intermediate rather than strongly elevated (Madagascar +0.43, Mozambique +0.41, Malawi +0.41, and Comoros +0.43). Conversely, Gabon ranked among the Bayesian top five (+1.48), but scored near zero in the NN model (+0.05), while remaining positively elevated in the Hybrid model (+1.17).

#### 3.4.2. Coldspot agreement

The three models showed stronger agreement for low-risk countries than for hotspot rankings. Kuwait, Lebanon, Jordan, Saudi Arabia, and Georgia all appeared in the bottom five of both the NN and Hybrid models. Among these, Lebanon was the only country that also appeared in the Bayesian bottom five. The Bayesian bottom five instead included the United States, Egypt, the Cook Islands, and Palestine. Overall, the Middle East showed the most consistent cross-model coldspot signal, especially in the NN and Hybrid maps.

#### 3.4.3. Summary

Taken together, the three models converged on two main findings: (i) SSA was the only clearly dominant high-risk macro-region for mosquito-borne disease co-occurrence; and (ii) the Middle East consistently contained several of the strongest coldspots, particularly in the NN and Hybrid models.

The principal disagreement lay in internal ranking within SSA, especially for several East and Southern African countries, which were strongly elevated by the NN model but remained near zero or only modestly elevated in the Bayesian and Hybrid models, and for Gabon, which was strongly prioritised by the Bayesian model but not by the NN model.

### 3.5. Stability map

Detailed data are presented in Supplementary Table S1.

The cross-model stability classification (see Table 1):

**Table 1.** cross-model stability classification.

| **Tier** | **$S_i$** | **Count** | **Proportion** |
| --- | --- | --- | --- |
| Non-hotspot | 0 | 99 | 69.7% |
| Low | 1 | 7 | 4.9% |
| Moderate | 2 | 7 | 4.9% |
| **Consensus** | **3** | **29** | **20.4%** |

**All 29 consensus hotspot countries (*S***_***i***_ **= 3)** were located in sub-Saharan Africa, spanning West Africa (Liberia, Sierra Leone, Guinea, Guinea-Bissau, Nigeria, Ghana, Côte d’Ivoire, Senegal, Gambia, Mali, Burkina Faso, Niger, Togo, and Benin), Central Africa (Cameroon, São Tomé and Príncipe, Congo, Democratic Republic of the Congo, Central African Republic, Chad, and Equatorial Guinea), East Africa (Ethiopia, Uganda, Burundi, Eritrea, Somalia, South Sudan, and Tanzania), and Southern Africa (Angola).

#### Moderate tier (*S*_*i*_ = 2; 7 countries)

The seven watch-list countries were Sudan, Gabon, Rwanda, Kenya, Mauritania, Zambia, and Suriname. Six were in sub-Saharan Africa, whereas Suriname was the only non-sub-Saharan African country in the *S*_*i*_ ≥ 2 group (Bayesian +1.17, NN +0.43, Hybrid +0.85) (see Figure 4 for more details).

**Figure 4.**
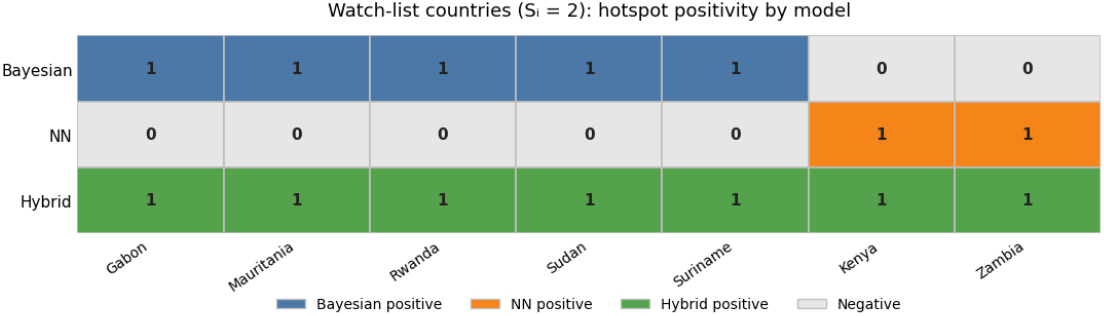
Cross-model hotspot positivity matrix for watch-list countries (Si=2) (Alt text: Matrix showing hotspot positivity across three models for the seven watch-list countries.)

#### Low tier (= 1; 7 countries)

Peru and Venezuela were Bayesian-only countries, whereas Comoros, Madagascar, Mozambique, Malawi, and Eswatini were NN-only countries. The Bayesian-only countries were located in South America, whereas the NN-only countries were located in East and Southern Africa. The geographic distribution of cross-model stability tiers is shown Figure 5

**Figure 5.**
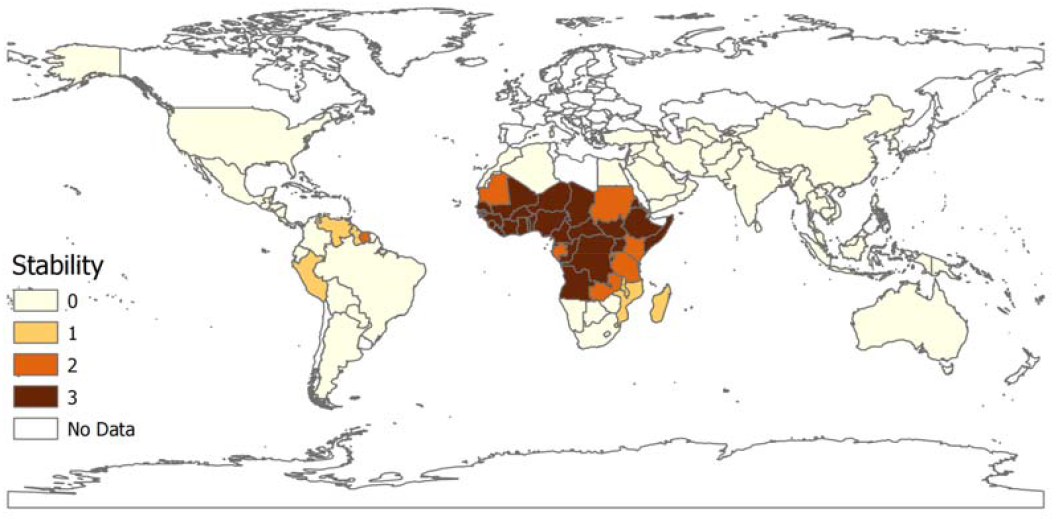
Cross-model stability classification map of shared mosquito-borne disease hotspots (Alt text: World map showing cross-model stability classification of shared mosquito-borne disease hotspots.)

## 4. Discussion

### 4.1. Main finding

This study showed that shared hotspots of dengue, malaria, and yellow fever were concentrated overwhelmingly in sub-Saharan Africa, with 29 countries reaching consensus hotspot status across all three models. Cross-model agreement was strongest for a contiguous tropical African hotspot belt, while only a small number of countries fell into intermediate tiers, indicating a relatively clear separation between the main high-burden core and surrounding fringe areas.

### 4.2. Comparison with recent global risk-oriented studies

A recent global study of Aedes-borne arboviruses used a multi-disease ecological niche model with a nested surveillance model to estimate environmental suitability and surveillance-adjusted risk, highlighting broad areas of potential vulnerability across South America, Southeast Asia, the Indian subcontinent, and parts of West and Central Africa^6^. In contrast, our study addressed a different question by focusing on realised country-level burden rather than potential suitability. Using observed epidemiological data for dengue, malaria, and yellow fever from 142 countries, we identified a much more concentrated pattern of robust shared hotspots, almost entirely confined to sub-Saharan Africa.

This contrast is especially clear in South America. In the recent suitability-oriented study, South America emerged as one of the major environmentally suitable regions for Aedes-borne arboviruses. However, within our joint three-disease framework, South America did not form a stable core of shared burden hotspots: Suriname was the only country reaching the moderate-agreement tier, while Peru and Venezuela appeared only in the low-agreement tier. This discrepancy suggests that environmental suitability does not necessarily translate into sustained realised multi-disease burden at the country level.

At the same time, both studies converge in identifying sub-Saharan Africa as a region of major concern. The recent global suitability study highlighted West and Central Africa as important environmentally suitable areas, while our results went a step further by showing that sub-Saharan Africa was not only suitable, but also the only region where robust shared burden hotspots consistently emerged across Bayesian, neural-network, and hybrid models. In this sense, our findings move from potential vulnerability to realised burden concentration.

An additional reason for the difference in spatial pattern is that our study included malaria alongside dengue and yellow fever, thereby shifting the analytical focus from Aedes-borne viral suitability to broader integrated mosquito-borne disease burden. From a public health perspective, this makes our hotspot classification more directly relevant for identifying geographic weak points in vector control, surveillance, and health-system preparedness. External validation against mortality burden further supports that these identified hotspots reflect not only co-occurrence of incidence, but also deeper weaknesses in prevention and control capacity.

### 4.3. Public health implications

A key public health implication of this study is that a shared hotspot across three mosquito-borne diseases should not be interpreted as equivalent to the hotspot pattern of any single disease. Instead, cross-disease co-occurrence may indicate a broader and more structural form of public health vulnerability, in which multiple mosquito-borne diseases converge within the same geographic settings. Under this interpretation, the identified hotspot countries are important not simply because they have high burden for one disease, but because they may represent settings where vector control, surveillance, prevention, and health-system response remain jointly constrained across diseases.

The identification of 29 SSA countries as consensus hotspots for the co-occurrence of dengue, malaria, and yellow fever has direct implications for vector control strategy. All three diseases are transmitted by mosquitoes (primarily by Anopheles mosquitoes for malaria; Aedes aegypti for dengue and yellow fever), and the geographic overlap of their high-burden zones suggests that integrated vector management (IVM) approaches targeting multiple vector species simultaneously may be more cost-effective in these countries than disease-specific interventions alone. The consensus hotspot map (Figure 5) can serve as a prioritisation tool for allocating IVM resources, identifying the 29 countries where cross-disease benefits of vector control are most likely to be realised.

The stability framework also identifies the *S*_*i*_ = 2tier (7 countries) as a watch list, representing countries that lie near the threshold of consensus hotspot status and may shift into full consensus under changing environmental, demographic, or surveillance conditions. From a policy perspective, these countries may be especially important for early warning and targeted surveillance, because intensified vector surveillance, sentinel monitoring, and earlier preventive action in such settings could help prevent consolidation into a more stable multi-disease hotspot profile.

### 4.4. Conclusion

In conclusion, this study identified a robust and highly concentrated pattern of shared mosquito-borne disease hotspots, centred overwhelmingly in sub-Saharan Africa. By moving beyond potential suitability and focusing on empirically observed shared burden across dengue, malaria, and yellow fever, the study provides a more actionable basis for identifying countries where vector control, surveillance, and health-system preparedness may be jointly constrained. These findings support more integrated prioritisation of mosquito-borne disease prevention and control in high-burden settings.

### 4.5. Limitation

First, the study relies on GBD modelled estimates of disease incidence and mortality, whose accuracy may vary across countries, particularly where national surveillance systems are weaker. Second, the analysis is ecological (country-level) and cannot identify within-country heterogeneity. Third, although the models included seven country-level covariates (temperature, humidity, rainfall, urbanisation rate, population density, physician density, and UHC index), these were annual national averages and did not capture subnational spatial variation or seasonal dynamics. Fourth, the external construct validation used cumulative mortality aggregated over the entire study period (1990–2023) rather than a strict out-of-sample window. Fifth, although the Stage-1 neural network exhibited signs of overfitting (training MSE ≈ 3.0 versus validation MSE ≈ 8.3), the primary outputs used for cross-model comparison were the country-level latent embeddings rather than the predicted incidence values. The strong external validation correlation (Pearson ρ = 0.83, p < 0.001) and the concordance between NN-derived shared scores and the other two models for consensus hotspot classification suggest that the latent representations retained meaningful geographic signal despite imperfect predictive fit on held-out years. Additionally, although the minimum ESS_bulk for the Hybrid model reached 229, falling below the conventional threshold of 400, this was concentrated in a small subset of parameters rather than being widespread; the corresponding R-hat values remained below 1.01 throughout, and the pattern was interpreted as modest local sampling inefficiency rather than substantive non-convergence.

## Data Availability

The data used in this study were obtained from publicly available, aggregated, and de-identified sources. The original data sources are described in the manuscript. The processed data and related analysis materials are currently stored in a Google Drive folder and are available at the link provided below.

https://docs.google.com/document/d/1VRH_FtvjntejsHA6gCzsBTRdkbcf8sbP/edit

## Author statements

### Authors’ contributions

Qiancheng Ma: Conceptualization, Formal analysis, Methodology, Visualization, Writing – original draft. Zhang Tianzhen: Validation, Writing – review & editing. All authors read and approved the final manuscript.

## Acknowledgements (if required)

The author acknowledges the Institute for Health Metrics and Evaluation and collaborators of the Global Burden of Disease Study 2023 for making the data publicly available.

## Funding

No specific funding was received for this work.

## Conflicts of interest

None declared.

## Ethical approval

Ethical approval was not required for this study because it used publicly available, anonymised secondary data and did not involve direct contact with human participants.

## Data availability statement

The data sources used in this study and the data generated during the analytical process are currently stored in a <u>private anonymized repository</u> to facilitate peer review and to minimize the risk of data leakage. Upon acceptance of the manuscript, the authors are willing to deposit the relevant data in an appropriate public repository.

## Declaration of generative AI and AI-assisted technologies in the manuscript preparation process

During the preparation of this work, the authors used ChatGPT to assist with translation and language editing in order to improve clarity and readability. The authors also used Claude to assist with code development as part of the research workflow. After using these tools, the authors reviewed and edited the content and code as needed and take full responsibility for the content of the publication.

## Notes

### Competing Interest Statement

The authors have declared no competing interest.

