## Supplementary Text 1 for "Global co-hotspots of dengue, malaria, and yellow fever: a comparative spatiotemporal analysis across 142 countries"

**Supplementary Material Text 1**

### 1. Notation and panel structure

Let $i = 1, \ldots, N$index countries (*N* = 142),$t = 1, \ldots, T$ index years (*T* = 34; 1990–2023), and *d* = 1, 2, 3 index diseases (dengue, malaria, yellow fever). The training window is$t\in\mathcal{T}_{t}r=1990,\ldots,TRAIN_{E}ND$; the out-of-sample window is$t\in\mathcal{T}_{t}e$. Observed case counts are $y_{i,d,t}\in Z_{\geq0}$; the population offset is $E_{i,t}=max\left( pop_{i,t},1 \right)$; the log-offset is$logE_{i,t}$. Country-level covariates are stacked into a tensor$X\in R^{N\times T\times K}$, where *K* is the number of covariates; within-training standardisation was applied per covariate to prevent validation-year leakage.

The country adjacency graph $A\in{0,1}^{N\times N}$ encodes land-border and short-sea neighbours; its edge list is$\left( e_{i},e_{j} \right)$ and the ICAR scaling factor *τ* was computed following Riebler et al. (2016) to make the marginal variance of the ICAR field invariant to the graph size(Riebler et al., 2016).

### 2. Model 1: Bayesian shared-component model

#### 2.1. Main model(Knorr-Held & Best, 2001)

$$y_{i,d,t}\sim\text{NegBin}(\mu_{i,d,t},\alpha_{d}^{\left( disp \right)})$$

$$log\mu_{i,d,t}=logE_{i,t}+\alpha_{d}+\sum_{k=1}^{K} \gamma_{d,k}x_{i,t,k}+\lambda_{d}\cdot s_{i}+u_{d,i}+\theta_{d,t}$$

##### 2.1.1. Component definitions

Shared spatial field (scaled ICAR):

$$s_{i}=\frac{\sigma_{s}}{\sqrt{\tau}}\cdot\varphi_{i},\varphi\sim\mathrm{ICAR}(A)$$

Disease-specific temporal trend (RW(1))(Mahaki et al., 2018):

$$\theta_{d,t}=\theta_{d,t-1}+\sigma_{\mathrm{rw},d}\cdot\eta_{d,t},\theta_{d,1}=0$$

##### 2.1.2. Symbol definitions

$y_{i,d,t}$— observed case count in country *i*, disease *d*, year *t*.

$\mu_{i,d,t}$ — expected count; mean of the Negative Binomial.

$\alpha_{d}^{NB}$ — disease-specific Negative Binomial dispersion (overdispersion) parameter.

$E_{i,t}$— population offset (so the model estimates rates, not absolute counts).

$\alpha_{d}$— disease-specific log-scale intercept.

$x_{i,t,k}$ – standardised value of covariate k for country i in year t.

$\gamma_{d,k}$— disease-specific coefficient of covariate *k*.

$K$ — number of covariates.

$s_{i}$— shared spatial component (primary quantity of interest), common to all three diseases.

$\lambda_{d}$— disease-specific loading on the shared spatial component.

$u_{d,i}$ — disease-specific residual spatial noise.

$\theta_{d,t}$ — disease-specific RW(1) temporal trend, centred to sum to zero.

$\varphi_{i}$— raw ICAR latent variable, centred.

*A* — country adjacency matrix (land-border and short-sea neighbours).

*τ* — ICAR scaling factor (Riebler et al. 2016).

$\sigma_{s},\sigma_{sd},\sigma_{rw,d}$ — scales of shared field, residual spatial noise, RW(1) innovations.

$\eta_{d,t}$ — standard-normal RW(1) innovation.

*N* = 142 (countries), *D* = 3 (diseases), $T_{t}r$(training years).

#### 2.2. Shared-score extraction (Model 1)

The primary reported output from Model 1 was the continuous Bayesian shared score, defined as:

$$\bar{s}_{i}^{\mathrm{Bayes}}=E[s_{i}\mid y]$$

**Symbols in the formula:**

- $s_{i}$— the shared spatial component for country *i* from the linear predictor in 2.1 (Model 1 main formula); a scalar random variable under the posterior distribution.
- *y* — the full set of observed case counts $y_{i,d,t}$ across all countries, diseases, and training years, serving as the conditioning data.
- $E[\cdot| y]$ — the posterior-mean operator: the average of $s_{i}$over all posterior draws after MCMC convergence.
- $s̄_{i}^{Bayes}$— the resulting country-level point estimate of the shared spatial position; the bar notation indicates "posterior mean".

3. **Bayesian implementation**

All unknown parameters and latent effects in Model 1 were assigned prior distributions and estimated jointly under a Bayesian framework. Thus, inference was based on the joint posterior distribution

$$\boldsymbol{p(}\boldsymbol{\Theta}\boldsymbol{\mid y)\propto p(y\mid}\boldsymbol{\Theta}\boldsymbol{)}\text{ }\boldsymbol{p(}\boldsymbol{\Theta}\boldsymbol{),}$$

where *p(y | Θ)* is the Negative Binomial likelihood defined in Section 2.1, and *p(Θ)* denotes the product of the prior distributions over the shared spatial field, disease-specific spatial effects, temporal effects, regression coefficients, intercepts, loadings, and dispersion parameters. Posterior inference was performed using the No-U-Turn Sampler (NUTS) with 4 chains, 4,000 warm-up iterations and 3,000 post-warm-up draws per chain (12,000 posterior draws in total), and a target acceptance rate of 0.95. Detailed prior specifications are listed in Table 1.

#### 3.1. MCMC sampler settings

Software: PyMC 5.28.2 with PyTensor 2.38.2 (MKL-linked); NUTS sampler.

Draws per chain: 3000

Tuning per chain: 4000

Chains: 4; cores: 4 (parallel).

target_accept: 0.95.

max_treedepth: 10.

Random seed: fixed.

Log-likelihood stored for WAIC/LOO.

Table 1 Model parameters

| Parameter | Specification |
| --- | --- |
| $\alpha_{d}$ | *N*(0, 2²) |
| $\gamma_{d,k}$ | *N*(0, 1²) |
| $\lambda_{d=dengue}$ | fixed at 1 |
| $\lambda_{d\neq dengue}$ | Half-Normal(0, 1²) |
| $s_{i}$ | Scaled ICAR, scale $\sigma_{s}$~ Half-Normal(0, 1²) |
| $u_{d,i}$ | $N\left( 0,*\sigma_{sd}*^{2} \right),with\sigma_{sd}\sim Half-Normal\left( 0,{0.3}^{2} \right)$ |
| $\theta_{d,t}$ | $RW\left( 1 \right),innovationscale\sigma_{rw,d}\sim Half-Normal\left( 0,{0.5}^{2} \right)$ |
| Likelihood | $NegativeBinomial,dispersion\alpha_{d}^{NB}\sim Gamma\left( 2,0.5 \right)$ |
| Sampler | PyMC 5.28.2, NUTS, 4 chains, target_accept = 0.95, max_treedepth = 10, fixed seed |

#### 3.2. Diagnostic checks

**Potential scale reduction factor (*R̂*)** — compares the variance within each MCMC chain to the variance between chains(Team, 2025b). **Purpose**: to check whether the four independent chains have converged to the same posterior distribution. If *R̂* is appreciably greater than 1, the chains have not mixed and the posterior summaries are unreliable. **Threshold**: *R̂* < 1.01 for all parameters(Team, 2025b).

**Effective sample size, bulk and tail (ESS_bulk, ESS_tail)** — estimates of the number of effectively independent draws from the posterior, accounting for autocorrelation within chains; ESS_bulk concerns the centre of the posterior (means, medians), ESS_tail concerns the extremes (credible-interval endpoints)(Team, 2025b). **Purpose**: to check that the Monte Carlo error on posterior summaries is small enough to make point estimates and intervals trustworthy. **Threshold**: ESS_bulk > 400 and ESS_tail > 400.

**Divergent transitions** — transitions flagged by the NUTS sampler when the leapfrog integrator fails in a region of high posterior curvature(Team, 2025a). **Purpose**: to detect pathological posterior geometry (often associated with funnels in hierarchical models) that would bias posterior exploration. **Threshold**: divergences should be zero after the tuning phase; a non-zero count is reported transparently and interpreted as a warning that specific posterior regions may be under-sampled.

**Posterior predictive check on *log(1 + y)* scale** — draws replicated data $y^{rep}$ from the fitted posterior predictive distribution and compares them to the observed *y* on the *log(1 + y)* scale(Team, 2025c). **Purpose**: to check whether the model can reproduce the observed count distribution. If PPC means systematically depart from the observed values, the likelihood or linear predictor is mis-specified. **Evaluation**: observed-vs-PPC-mean scatter with *y = x* reference line; systematic off-diagonal patterns flag fit problems.

**Within-family WAIC and LOO elpd** — WAIC and LOO were examined jointly within the NB family. **Purpose:** to assess predictive fit, effective complexity, and the stability of model evaluation. **Key quantities:** the main quantities of interest were $elpd_{waic}$and $elpd_{loo}$, with higher (less negative) values indicating better expected out-of-sample predictive performance under the same likelihood and observation scale(Team, 2025c). Agreement between WAIC and LOO was also examined; in this study, closer values were interpreted as indicating more stable model evaluation.

### 4. Model 2: Neural network (LSTM + spatial lag)

#### 4.1. Main model(Hochreiter & Schmidhuber, 1997)

$$\mathbf{h}_{i,t}=\mathrm{LSTM}\text{ }\left( {\tilde{\mathbf{x}}}_{i,1:T} ; \text{ }\Theta_{\mathrm{lstm}} \right)_{t}$$

$$\hat{y}_{i,d,t}^{\log}=f_{d}\text{ }\left( [\mathbf{h}_{i,t};\text{ }\mathbf{z}_{i}];\text{ }\Theta_{d} \right), d=1,2,3$$

##### 4.1.1. Component definitions

**Input with spatial lag:**

The neural network input was augmented with the neighbour-averaged covariates Wx alongside the own-country covariates x, following the spatial-lag-of-X (SLX) specification of Halleck Vega and Elhorst (2015), so that spatial dependence could be encoded on the input side rather than through a separate spatial random effect(Vega, 2015).

$${\tilde{\mathbf{x}}}_{i,t}=\left[ \text{ }\mathbf{x}_{i,t}^{\mathrm{std}}\text{ };\text{ }(W\mathbf{x}^{\mathrm{std}})_{i,t}\text{ } \right], W=D^{-1}A$$

##### 4.1.2. Country-level latent embedding (time-averaged encoder output):

$$\mathbf{z}_{i}=\mathrm{Enc}\text{ }\text{ }\left( \frac{1}{T}\sum_{t=1}^{T} \mathbf{h}_{i,t} ; \text{ }\Theta_{\mathrm{enc}} \right)$$

##### 4.1.3. Training loss (MSE on log-rate):

$$\mathcal{L=}\frac{1}{N\cdot\mid\mathcal{T}_{\mathrm{tr}}\mid\cdot D}\sum_{i,t,d} \left( \hat{y}_{i,d,t}^{\log} - y_{i,d,t}^{\log} \right)^{2}$$

#### 4.2. Symbol definitions

$x_{i,t}^{std}$ — standardised covariate vector.

$W=D^{-1}A$ — row-normalised adjacency matrix; $\left( Wx \right)_{i,t}$ is the neighbour-averaged covariate.

$x̃_{i,t}$ — LSTM input: concatenation of own and spatial-lag covariates.

$h_{i,t}\in\mathbb{R}^{H}$ — LSTM hidden state; *H* = hidden size.

$\Theta_{lstm},\Theta_{enc},\Theta_{d}$— trainable weights of LSTM, encoder MLP, and per-disease heads.

$z_{i}\in\mathbb{R}^{K_{z}}$ — country-level latent embedding; $K_{z}$ = embedding dimension.

$f_{d}$— per-disease prediction head (small MLP).

$ŷ_{i,d,t}^{log}$— predicted log-incidence rate.

$y_{i,d,t}^{log}=log\left( y_{i,d,t}/E_{i,t}\times{10}^{5}+1 \right)$ — training target.

$\mathcal{T}_{t}r$ — set of training years.

$\mathcal{L}$ — mean squared error loss.

#### 4.3. Model parameters

| **Component** | **Specification** |
| --- | --- |
| Input | standardised country-level covariates concatenated with row-normalised spatial-lag covariates (Wx,W=row-normalised adjacency matrix) |
| LSTM | 2 stacked layers, hidden size $H = 64$ |
| Encoder (produces (z_i)) | Linear$\left( H \to32 \right)$ $\to$ReLU $\to$Linear$\left( 32 \to K_{z} \right)$ $\to$ReLU; $K_{z}=16$ |
| Per-disease head (f_d) | Linear$\left( H+K_{z}\to32 \right)$ $\to$ReLU $\to$Linear$\left( 32 \to1 \right)$; three heads in parallel |
| Training target | log(cases/pop×10^5^+1) |
| Loss | mean squared error |
| Optimiser | Adam, $lr=1\times{10}^{-3}$, weight decay = $1\times{10}^{-4}$ |
| Gradient clipping | max-norm (1.0) |
| LR scheduler | ReduceLROnPlateau ($\text{patience}=10,;\text{factor}=0.5$) |
| Validation split | last 20% of training years |
| Early stopping | patience = 80 on validation MSE; best checkpoint restored |
| Seeds | fixed for torch and numpy |
| Device | CUDA if available, else CPU |

#### 4.4. Shared-score extraction

$$s_{i}^{\mathrm{NN}}=\mathrm{sign}\text{-}\mathrm{oriented}\text{ }\text{ }\left( \mathrm{PC}1\text{ }\text{ }\left( \{ \mathbf{z}_{i}{\}}_{i=1}^{N} \right)_{i} , \text{ }a \right)$$

PC1 is the first principal component of the embedding matrix *Z* (Jolliffe & Cadima, 2016);Sign-oriented against the disease anchor *a*.

#### 4.5. Diagnostic checks

**Training and validation MSE trajectories per epoch(Prechelt, 2002)** — the MSE of predicted vs. observed log-rate evaluated on the training fold and on the held-out 20% validation fold, plotted against epoch. **Purpose**: to confirm that the network is learning (training loss decreases) and to detect overfitting (validation loss stops decreasing or starts to rise while training loss keeps decreasing). **Evaluation**: visual inspection of the two curves.

**Early-stopping epoch(Prechelt, 2002)** — the epoch at which training was halted because the validation MSE stopped improving for NN_PATIENCE consecutive epochs. **Purpose**: to document the actual training duration and confirm that early stopping activated (rather than hitting the epoch cap), which is an indirect indicator that the chosen patience and maximum-epoch settings were appropriate. **Reported value**: a single integer per run.

**Out-of-sample test MSE** — MSE of predicted vs. observed log-rate on years after TRAIN_END, which the model has never seen. **Purpose**: to assess genuine out-of-sample generalisation. This is the NN's analogue of Bayesian WAIC/LOO — both measure predictive performance on unseen data, but test MSE does so directly on held-out years rather than by approximation(Bergmeir & Benítez, 2012). **Evaluation**: absolute value compared against training/validation MSE (large test-vs-train gap indicates distribution shift or overfitting).

**Predicted-vs-observed scatter on log-rate scale, per disease** — scatter plot for each disease separately, with *y = x* reference line. **Purpose**: to check whether the model systematically under- or over-predicts certain ranges (e.g., low-incidence countries) and whether performance is comparable across diseases. **Evaluation**: visual; systematic curvature or fan-shaped residuals flag mis-specification of the head architecture or loss scale.

### 5. Model 3: Two-stage Hybrid

To our knowledge, however, this specific combination — an LSTM country-level embedding fed into a Bayesian observation layer with ICAR shared-component spatial structure, disease-specific RW(1) temporal trends, and cross-disease shared loadings — has not been applied to multi-disease hotspot identification for mosquito-borne diseases.

The Hybrid model uses Model 2 as a feature extractor. The neural network passes a single country-level embedding matrix$Z\in R^{N\times K_{z}}$ to the Bayesian layer, where each row $z_{i}$is a fixed, learned summary of country i. This embedding carries the non-linear covariate interactions, time-averaged country-level signal, and neighbour context that the NN has learned from the panel. The Bayesian layer takes Z as fixed input and adds what the NN cannot provide on its own: an ICAR-structured shared spatial field$s_{i}$, disease-specific RW(1) temporal trends $\theta_{d,t}$, and full posterior uncertainty for all quantities. The two stages are deliberately separated (no gradients flow back into the NN) to avoid identifiability conflicts observed when they were estimated jointly.

#### 5.1. Main formula

$$y_{i,d,t}^{\log}\mathcal{\sim N(}\mu_{i,d,t},\sigma_{\mathrm{obs},d}^{2})$$

$$\mu_{i,d,t}=\alpha_{d}+\sum_{k=1}^{K_{z}} \beta_{d,k}\text{ }z_{i,k}+\lambda_{d}\cdot s_{i}+\theta_{d,t}$$

#### 5.2. Symbols in the main formula

$\mu_{i,d,t}$ — mean log-incidence rate for country *i*, disease *d*, year *t*.

$\alpha_{d}$— disease-specific intercept.

$\beta_{d,k}$— disease-specific coefficient on the *k*-th NN latent feature.

$z_{i,k}$— *k*-th component of the fixed NN country embedding (from Model 2).

$\lambda_{d}$ — disease-specific loading on the shared spatial component.

$s_{i}$— shared spatial component.

$\theta_{d,t}$ — disease-specific RW(1) temporal trend.

#### 5.3. Bayesian implementation

Model 3 was estimated in two stages. In Stage 1, the neural network was trained to learn fixed country-level latent embeddings $z_{i}$. In Stage 2, these embeddings were treated as fixed covariates in a Bayesian observation layer, where the shared spatial field, disease-specific temporal effects, regression coefficients, loadings, and observation-level variance parameters were assigned prior distributions and inferred jointly from the posterior distribution

$$p(\Theta_{\text{Bayes}}\mid y,z)\propto p(y\mid\Theta_{\text{Bayes}},z)\text{ }p(\Theta_{\text{Bayes}}),$$

where $z$denotes the fixed NN embeddings imported from Model 2 and treated as observed inputs in Stage 2. Posterior inference was performed using NUTS, with detailed prior settings listed in Table 2.

Table 2 Model parameters

| Parameter | Specification |
| --- | --- |
| Stage 1 | NN embeddings $Z=z_{i}$ imported from Model 2, treated as fixed data (no gradients) |
| $\alpha_{d}$ | *N*(0, 1²) |
| $\beta_{d,k}$ | *N*(0, 0.5²) |
| $\lambda_{d=dengue}$ | fixed at 1 |
| $\lambda_{d\neq dengue}$ | LogNormal(0, 0.35²) |
| $s_{i}$ | scaled ICAR, scale $\sigma_{s}$~ Half-Normal(0, 0.5²) |
| $\theta_{d,t}$ | RW(1), innovation scale $\sigma_{rw,d}$ ~ Half-Normal(0, 0.5²) |
| Likelihood | Gaussian on log-rate scale, $\sigma_{obs,d}$~ Half-Normal(0, 0.5²) |
| Residual spatial noise $u_{d,i}$ | not included (removed due to identifiability conflict with $\beta_{d,k} z_{i,k}$ |
| Sampler | PyMC 5.28.2, NUTS, 4 chains, target_accept = 0.95, max_treedepth = 12, fixed seed |

#### 5.4. Shared-score extraction

The primary reported output from Model 3 was the continuous country-level Hybrid shared score. For each country $i$, the model first constructed a disease-specific Hybrid score vector by combining the posterior-mean shared spatial contribution and the posterior-mean NN-feature contribution across diseases:

$$\mathbf{s}_{i}^{\mathrm{Hybrid},\mathrm{by}\text{-}\mathrm{disease}}=\bar{\lambda}\text{ }\bar{s}_{i}+\bar{B}z_{i}$$

— The final reported country-level Hybrid shared score was then defined as the average of these disease-specific contributions:

$$\bar{s}_{i}^{\mathrm{Hybrid}}=\frac{1}{D}\mathbf{1}^{\top}\left( \bar{\lambda}\text{ }\bar{s}_{i} + \bar{B}z_{i} \right)$$

##### 5.4.1. Symbols in the formula

the final country-level Hybrid shared score for country $i$; a scalar obtained by averaging the disease-specific Hybrid contributions.

$\mathbf{s}_{i}^{\mathrm{Hybrid},\mathrm{by}\text{-}\mathrm{disease}}$— the disease-specific Hybrid score vector for country $i$, with length $D$.

$\bar{\lambda}$ — posterior-mean vector of disease-specific loadings on the shared spatial component, with $\lambda_{\text{dengue}}=1$fixed as the identifiability anchor.

$\bar{s}_{i}$ — posterior mean of the shared ICAR spatial field at country $i$.

$\bar{B}$— posterior-mean matrix of disease-specific coefficients on the NN latent features, with dimension $D\times K_{z}$.

$z_{i}$ — fixed NN country embedding for country $i$, imported unchanged from Model 2.

$D$*—* number of diseases included in the joint model.

#### 5.5. Diagnostic checks

**Potential scale reduction factor (*R̂*)** — same definition and purpose as in Model 1. **Threshold**: *R̂* < 1.01 for all parameters. Of particular interest here are the$\beta_{d,k}$ coefficients on NN latent features, because they are the channel through which Stage-1 representations enter the Bayesian layer; poor convergence on these parameters would indicate that the latent features are not well identified as covariates .

**Effective sample size, bulk and tail (ESS_bulk, ESS_tail)** — same definition and purpose as in Model 1. **Threshold**: target > 400 for both. In our runs the minimum observed ESS_bulk was 229, concentrated in a small number of *β* coefficients; we report this explicitly rather than masking it, and interpret it as a modest sampling inefficiency rather than a convergence failure (the corresponding *R̂* values remained below 1.01).

**Divergent transitions** — same definition and purpose as in Model 1. The max_treedepth was raised to 12 (from the default 10 used in Model 1) specifically to reduce tree-depth saturation observed in pilot runs, which had caused a small number of divergences. **Threshold**: zero post-tuning.

**Posterior predictive check on log-rate scale** — draws $y^{rep}$from the Gaussian observation layer and compares to the observed log-rate. **Purpose**: to check whether the Bayesian observation layer, conditional on the fixed NN representation *Z*, can reproduce the observed log-rate distribution. If the NN representation is carrying most of the signal, PPC fit here will be tighter than in Model 1. **Evaluation**: observed-vs-PPC-mean scatter; systematic curvature would suggest that linear combinations of $z_{i,k}$ are insufficient and that a non-linear transformation of *Z* might be needed.

**Within-family WAIC and LOO-IS elpd** — same method as in Model 1, applied to the Gaussian-on-log-rate family. **Purpose**: to compare variants of Model 3 (e.g., with or without $u_{d,i}$, different $K_{z}$) on a like-for-like basis. **Scope limitation**: not compared against Models 1 or 2 — against Model 1 because of likelihood and scale differences, against Model 2 because Model 2 has no explicit likelihood.

### 6. Score standardisation for cross-model comparison

The three models produce raw shared scores on different internal scales because they arise from distinct mathematical constructions.

**For Model 1 (Bayesian shared-component)**, the raw shared score was defined as the posterior mean of the shared ICAR spatial component, $\bar{s}_{i}^{\mathrm{Bayes}}=E[s_{i}\mid y]$. Because the ICAR specification is centred through the sum-to-zero constraint, these raw scores were naturally distributed around zero, with both positive and negative values.

**For Model 2 (Neural network)**, the raw shared score was defined as the first principal component of the country-level embedding matrix, $s_{i}^{\mathrm{NN}}=\mathrm{PC}1(\{z_{i}\})$. Because PCA is computed on centred features, the resulting raw scores were likewise centred near zero.

For Model 3 (Two-stage Hybrid), unlike the Bayesian ICAR score and the PCA-based NN score, the Hybrid score is not subject to an intrinsic zero-centering constraint. Consequently, its empirical raw-score distribution may be shifted away from zero. In our application, the Hybrid raw scores showed a clear positive shift (mean ≈ +2.6, range ≈ +0.03 to +5.94), indicating that direct comparison of raw values across models would be inappropriate.

To enable fair cross-model comparison, the raw score vector from each model was independently Z-standardised across the $N$countries:

$$z_{i}^{\left( m \right)}=\frac{s_{i}^{\left( m \right)}-\mathrm{mean}(s^{\left( m \right)})}{\mathrm{SD}(s^{\left( m \right)})},m\in\{\mathrm{Bayes},\mathrm{NN},\mathrm{Hybrid}\}$$

After standardisation, all three score vectors had mean $=0$and SD $=1$, placing them on a common scale and removing differences in centring and variance across models.

### 7. Cross-model stability rank

#### 7.1. Main formula

$$S_{i}=h_{i}^{\mathrm{Bayes}}+h_{i}^{\mathrm{NN}}+h_{i}^{\mathrm{Hybrid}}\in0,1,2,3$$

#### 7.2. Symbols in the main formula

$S_{i}$— stability rank of country *i*; the number of models that classify it as a hotspot.

$h_{i}^{\left( m \right)}$ ∈ {0, 1} — binary hotspot indicator under model *m*.

$$h_{i}^{\left( m \right)}=1\{z_{i}^{\left( m \right)}\geq P75\left( z^{\left( m \right)} \right)\}$$

where $z_{i}^{\left( m \right)}$is the country-level shared score from model $m$after sign-orientation against the disease anchor and standardisation to a Z-score across the 142 countries.

The cutoff $z_{i}^{\left( m \right)}\geq P75\text{ }\text{ }\left( z^{\left( m \right)} \right)$corresponds to “country $i$lies at or above the 75th percentile of the cross-country shared-score distribution under model $m$”.

Specifically: $h_{i}^{Bayes}=1$if the standardised posterior-mean Bayesian shared score $z_{i}^{Bayes}=\frac{\bar{s}_{i}^{Bayes}-\mathrm{mean}\text{ }\left( \bar{s}^{Bayes} \right)}{SD\text{ }\text{ }\left( \bar{s}^{Bayes} \right)}$satisfies $z_{i}^{Bayes}\geq P75\text{ }\text{ }\left( z^{Bayes} \right)$— i.e., country $i$lies at or above the 75th percentile of the Bayesian ICAR posterior shared field.

$h_{i}^{NN}=1$if the standardised first principal component of the NN country embeddings $z_{i}^{NN}$satisfies $z_{i}^{NN}\geq P75\text{ }\left( z^{NN} \right)$— i.e., country $i$lies at or above the 75th percentile along the dominant axis of NN-learned country-level variation.

$h_{i}^{Hybrid}=1$if the standardised Hybrid shared score $z_{i}^{Hybrid}$satisfies $z_{i}^{Hybrid}\geq P75\text{ }\text{ }\left( z^{Hybrid} \right)$— i.e., country $i$lies at or above the 75th percentile along the combined NN-feature plus ICAR-spatial axis.

#### 7.3. Method parameters

| Step | Specification |
| --- | --- |
| Sign orientation | each model's shared score $\tilde{s}_{i}^{\left( m \right)}$oriented against the disease anchor $a_{i}$(log of the summed three-disease incidence rate, averaged over training years) |
| Standardisation | sign-oriented scores Z-standardised across *N* = 142 countries within each model |
| Hotspot cutoff | $zi\left( m \right)\geq P75\left( z\left( m \right) \right)$ |
| Stability tiers | $S_{i}$= 0 non-hotspot; 1 low; 2 moderate; 3 consensus |

### 8. External validation

#### 8.1. Main formula

$$\rho^{\left( m \right)}=\mathrm{corr}\text{ }\text{ }\left( s^{\left( m \right)} , \text{ }M \right),m\in Bayes,\text{ }NN,\text{ }Hybrid$$

Two correlation coefficients are reported separately:

$$\rho_{S}^{\left( m \right)}=\mathrm{Spearman}\text{ }\text{ }\left( s^{\left( m \right)} , \text{ }M \right),\rho_{P}^{\left( m \right)}=\mathrm{Pearson}\text{ }\left( s^{\left( m \right)} , \text{ }M \right)$$

#### 8.2. Symbols in the main formula

$\rho^{\left( m \right)}$ — correlation coefficient between model *m*'s shared score and the mortality anchor *M*. Reported as two complementary coefficients: **Spearman rank correlation** $\rho_{S}^{\left( m \right)}$measures how well model *m* preserves the country ranking along the mortality axis (sensitive to monotonic relationships but not to linearity); **Pearson linear correlation**$\rho_{P}^{\left( m \right)}$measures the strength of the linear association (sensitive to magnitude matching, not just rank).

$s^{\left( m \right)}$— sign-oriented shared score from model *m* (before Z-standardisation).

*M* — country-level log-mortality anchor constructed from **GBD death counts for the same three mosquito-borne diseases studied in this work (dengue, malaria, and yellow fever)**. Aggregated cumulatively across all panel years (1990–2023).

#### 8.3. Method parameters

| Step | Specification |
| --- | --- |
| Mortality data source | GBD death counts for the same three mosquito-borne diseases used in model training (dengue, malaria, yellow fever) |
| Aggregation window | entire panel period (1990–2023); cumulative across all available years |
| Mortality anchor $M_{i}$ | $M_{i}$ *=* log( ($\Sigma_{d}\Sigma_{t}\mathrm{death}s_{i,d,t}$) / ($\Sigma_{t}\mathrm{po}p_{i,t}$) × 10⁵ + 1 ) — cumulative total deaths across three diseases and all years, divided by cumulative population, log-transformed with +1 offset |
| Sample size | *N* = 142 countries, identical across the three models |
| Correlation metrics | Spearman (rank-based) and Pearson (linear), both reported; two-sided p-values |
