## Supplementary Text 2 for "Global co-hotspots of dengue, malaria, and yellow fever: a comparative spatiotemporal analysis across 142 countries"

1. **Diagnostic checks (Model 1: Bayesian shared-component model)**

Posterior convergence was assessed using the potential scale reduction factor ($\hat{R}$) and effective sample size (ESS). All monitored parameters satisfied the predefined convergence criterion of $\hat{R}<1.01$. The maximum observed $\hat{R}$was 1.0043, while the minimum $ESS_{bulk}$and $ESS_{tail}$were 492.4 and 1050.2, respectively. These results indicate good chain mixing and adequate posterior sampling efficiency for all monitored parameters. These results indicate good chain mixing and adequate posterior sampling efficiency for all monitored parameters (see Figure S1)

No post-tuning divergent transitions were observed, indicating that the NUTS sampler explored the posterior geometry without obvious pathological behaviour.

Posterior predictive checks (PPC) on the $\log(1+y)$scale showed that replicated data from the fitted posterior predictive distribution broadly reproduced the observed count distribution. The observed-versus-PPC-mean scatter was concentrated around the $y=x$reference line without marked systematic departure, supporting adequate model fit (see Figure S2).

Within-family WAIC and LOO diagnostics were also broadly concordant. The estimated elpd_{WAIC}and elpd_{LOO}were -83193.81 and -83203.05, respectively, indicating stable internal predictive evaluation for the Bayesian model family.

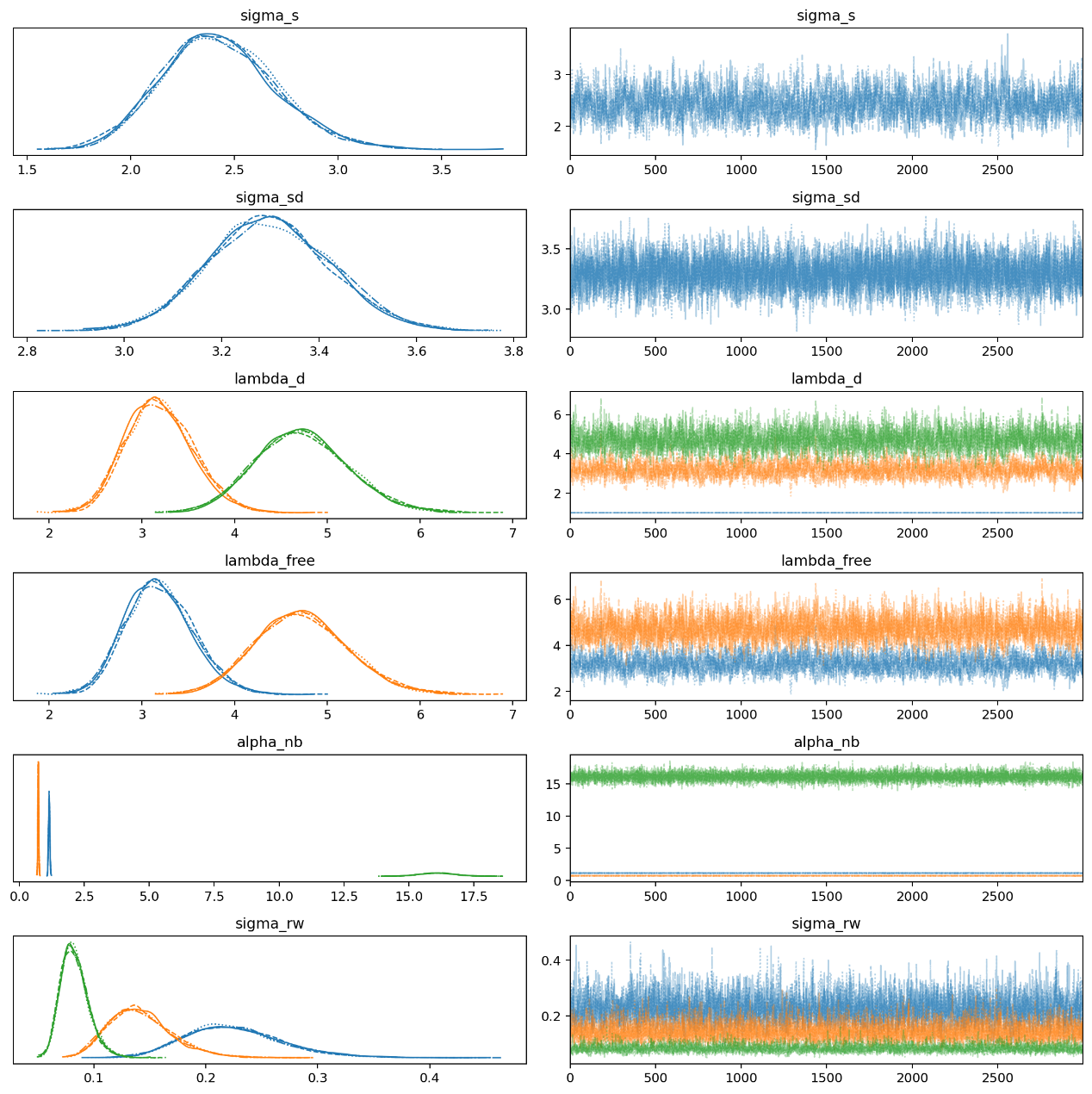

Figure S1 Posterior trace plots for selected parameters in Model 1.

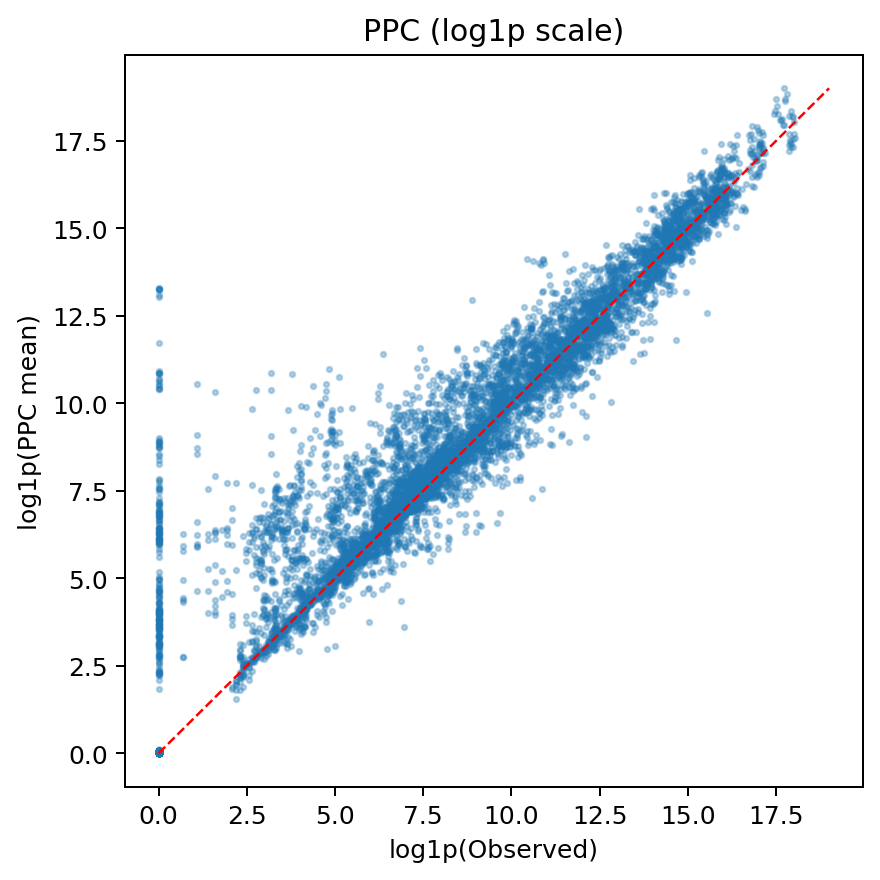

Figure S 2 Posterior predictive check for Model 1 on the \log\funcapply(1+y) scale.

2. **Diagnostic checks (Model 2: Neural network)**

Diagnostic evaluation for Model 2 focused on optimisation behaviour and out-of-sample predictive performance rather than MCMC-based convergence metrics. The model was trained under an explicit early-stopping rule, with a maximum of 1500 epochs and a patience of 80 epochs. After stopping, the best validation checkpoint was restored.

The loss trajectories showed substantial improvement in both training and validation MSE during optimisation. Training MSE decreased from approximately 21.0 at the start of training to about 3.0 at the end, while validation MSE decreased from about 18.6 to about 8.3. The validation curve entered a broad plateau after roughly 120 epochs, whereas the training curve continued to decline, indicating mild overfitting in the later stage of optimisation rather than optimisation failure(see Figure S3 for more datils).

Observed-versus-predicted plots on the log-rate scale showed positive correspondence between predictions and observations across all three diseases, confirming that the network captured broad cross-country variation. The predictive relationship was clearest for malaria, whereas dengue and yellow fever showed greater scatter. Across diseases, the prediction range was compressed relative to the observed range, with systematic underestimation at higher observed log-rates. This pattern suggests acceptable overall learning, but weaker fit in the upper tail of the distribution(see Figure S4).

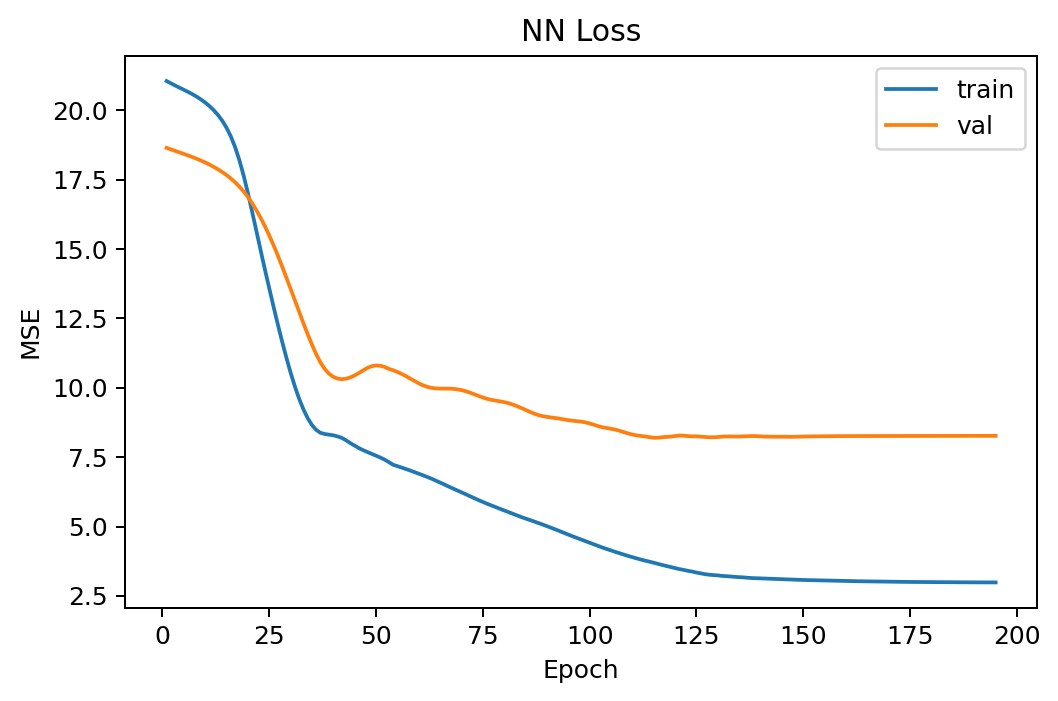

Figure S3Training and validation MSE trajectories for Model 2

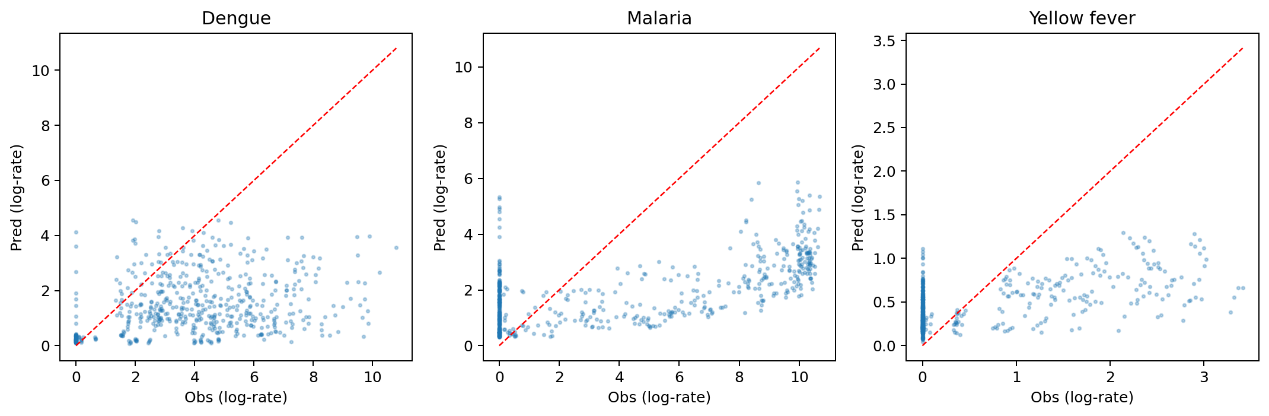

Figure S4 Observed-versus-predicted log-rate plots for Model 2 across the three diseases.

3. **Diagnostic checks (Model 3: Two-stage Hybrid)**

For Model 3, posterior convergence of the Bayesian observation layer was assessed using the same criteria as in Model 1. All monitored parameters satisfied the predefined threshold of $\hat{R}<1.01$, with a maximum observed $\hat{R}$of 1.0096. The minimum $ESS_{bulk}$and $ESS_{tail}$were 229.2 and 533.2, respectively. Although the minimum $ESS_{bulk}$fell below the target value of 400, these lower ESS values were concentrated in a small subset of parameters rather than being widespread across the model, and the corresponding $\hat{R}$values remained below 1.01. This pattern was therefore interpreted as modest local sampling inefficiency rather than substantive non-convergence. These results indicate good chain mixing and adequate posterior sampling efficiency for all monitored parameters (see Figure S5)

No post-tuning divergent transitions were retained in the final Hybrid fit. To reduce tree-depth saturation observed in pilot runs, max_treedepth was increased from 10 to 12 in the final sampling configuration.

Posterior predictive checks on the log-rate scale indicated acceptable fit of the Gaussian Bayesian observation layer conditional on the fixed NN representation $Z$. The observed-versus-PPC-mean scatter showed no strong systematic deviation, suggesting that the Hybrid observation layer could adequately reproduce the main features of the observed log-rate distribution(see Figure S6).

Within the Gaussian-on-log-rate Hybrid family, WAIC and LOO values were also broadly concordant. The estimated $elpd_{WAIC}$and $elpd_{LOO}$were -18687.02 and -18687.38, respectively, indicating stable internal predictive evaluation for Model 3.

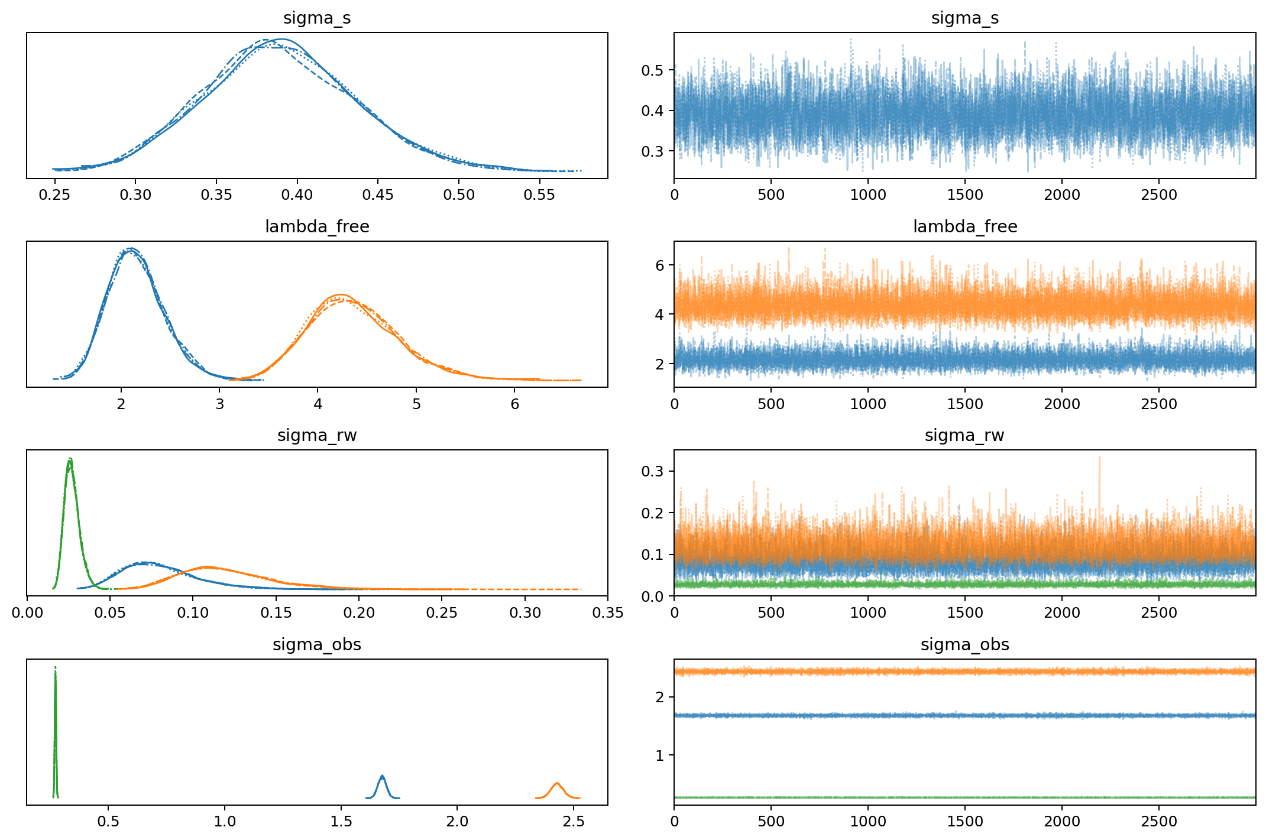

Figure S5 Trace plots for selected parameters in Model 3

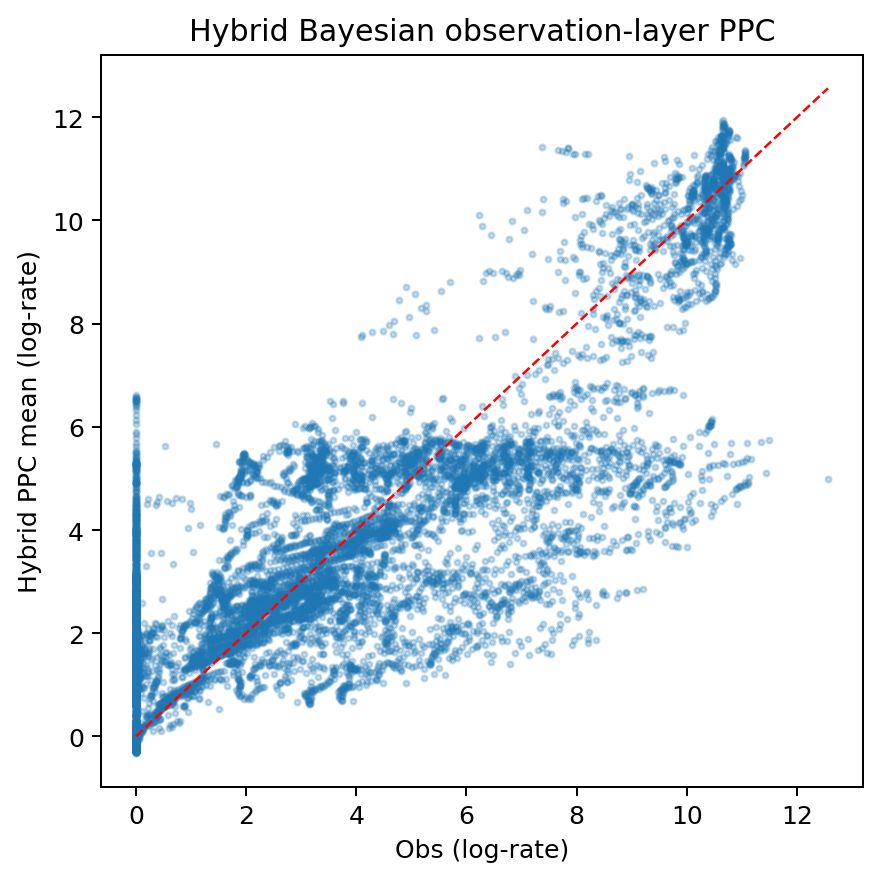

Figure S6 Posterior predictive check for Model 3 on the log-rate scale.
